# Study protocol for evaluating EEG-based predictive model for drowsiness measurement to reduce accident risk in active individuals

**DOI:** 10.1101/2025.04.26.25326496

**Authors:** Chloe Boitard, Khadijeh Sadatnejad, Christian Berthomier, Julien Coelho, Julie Lenoir, Patricia Sagaspe, Julien Mattei, Pierre Berthomier, Marie Brandwinder, Pierre Philip, Jean-Arthur Micoulaud Franchi, Jacques Taillard

## Abstract

Voluntary behaviors and socio-economic factors, such as social jetlag and shift work, can lead to insufficient or disrupted sleep, resulting in drowsiness in active individuals. In occupational and driving contexts, drowsiness poses a serious safety risk by impairing alertness, slowing reaction times, and increasing the likelihood of accidents. Developing predictive, automatic and easy to implement tools for drowsiness detection is essential in high-risk environments where sustained vigilance is critical. This study aims to validate a practical and predictive method for assessing drowsiness using automated analysis of a limited number of electroencephalogram (EEG) channels. Designed as single-center, non-randomized, single-group, this study will evaluate drowsiness and cognitive performance in forty healthy volunteers exposed to two sleep deprivation conditions simulating real-world occupational scenarios. The primary outcome will be the Objective Sleepiness Scale (OSS) and its automated analysis, with a focus on its ability to measure objective wakefulness as assessed by the Maintenance of Wakefulness Test (MWT). Secondary outcomes will include multimodal resting-state EEG markers, subjective and objective sleepiness measures, performance on a simulated driving task, attention, executive function and vigilance assessments, as well as sleep quality, sleep quantity, and mind-wandering. The influence of sociodemographic and clinical variables on drowsiness measurement and prediction will also be systematically examined. By validating these novel EEG-based measures, this study aims to lay the groundwork for proactive drowsiness management strategies in occupational, transportation, and clinical settings.

## 1. Introduction

Sleepiness is a physiological and behavioral “need state” or “need for sleep”. It plays a key role in the regulation of the sleep/wake cycle, especially in triggering sleep onset at the usual bedtime or during sleep deprivation. Sleepiness facilitates the transition from wakefulness to sleep and hinders the transition from sleep to wakefulness. “Physiological sleepiness”[1], a.k.a. sleep drive, results from an imbalance between processes involved in the regulation of sleep and wake states. It is orchestrated both by the sleep homeostatic process, relying on the prior duration of wakefulness and the prior amount of sleep, and by the circadian rhythm, inducing a forbidden sleep zone at the end of the evening and an imposed sleep zone at the end of the night. Physiological sleepiness is modulated by inter-individual differences (chronotype, age, sex).

The manifest sleepiness [1] is the transformational effect of underlying physiological sleepiness on behavior, due to voluntary or involuntary sleep deprivation, circadian misalignment or pathologies inducing excessive daytime sleepiness (EDS). Manifest sleepiness is also called: drowsiness, hypo-arousal [2] or continuous non-imperative sleepiness [3] by other authors. We will here use the generic term drowsiness to qualify manifest sleepiness. Drowsiness is characterized by an inappropriate wake behavior and by a lasting inadequate level of arousal which induces inability to stay awake, difficulties in maintaining sustained attention and vigilance (i.e. brain fog) and affects judgment, decision-making abilities and encourages risk-taking [4]. Drowsiness can thus induce functional consequences in terms of both disability and accident risk [5–7], in the same proportions as the effects of alcohol consumption [8]. Drowsiness also redirects attention to thought that are not related to the cognitive task realized [9]. This modification in attentional orientation, known as mind wandering, also impacts cognitive performance [9,10]. Finally, introspective sleepiness [1] concerns the individual’s self-assessment of its internal sleepiness state. Manifest and introspective measures are linked because they both reflect underlying sleep pressure and drowsiness, and therefore may stem from the same underlying drive state [11]. However, drowsy people often fail to accurately assess their level of drowsiness, and therefore overestimate their ability to make important decisions and perform complex tasks [12], despite their decreased cognitive abilities, which can lead to dangerous consequences. Identifying and measuring real-time drowsiness or predicting it in high-risk situations like working or driving is thus a significant public health challenge [13,14].

Drowsiness can result from voluntary behaviors or socio-economic factors that lead to insufficient or disrupted sleep [15], such as social jetlag [16] and shift work [17]. Drowsiness assessment over time is mainly based on brain activity measurements (electroencephalogram, or EEG) [18]. EEG provides a direct and objective window into neural correlates of alertness, unlike subjective measures or sporadic performance-based assessments. EEG data can reveal subtle transitions between wakefulness and drowsiness that may otherwise go undetected. Thus, the primary index of drowsiness, i.e. the ability to stay awake, is assessed during the Maintenance of Wakefulness Test (MWT) [19] an iterative electrophysiological test in which EEG and electrooculogram (EOG) analysis allow the determination of daytime sleep latency, identifying “global” daytime drowsiness. Moreover, behavioral consequences of drowsiness, i.e. impaired cognitive performances and reduced vigilance (characterized by degraded reaction time and increased commissions) are related to changes in EEG recordings, showing episodic microsleep intrusion, local sleep or wake-state instability [4,18]. Thereby, EEG analysis, considered as the “gold standard” for direct and continuous monitoring of sleepiness [18], represents the most promising way to detect drowsiness in safety-sensitive contexts such as transportation, healthcare, and industrial operations, where lapses in vigilance can have serious consequences.

The earliest EEG-based methods developed to assess spontaneous drowsiness in offline conditions include the Karolinska Drowsiness Score (KDS) [20]. This approach is designed to evaluate sleepiness- related impairments in performance, particularly in driving, by quantifying theta and alpha EEG activity and/or identifying specific slow rolling eye movements via electrooculogram (EOG), resulting in a continuous sleepiness score ranging from 0 (not sleepy) to 100 (very sleepy). Currently, numerous EEG- based systems and algorithms - both offline and online - have been proposed for the detection of drowsiness. These methods often rely on various frequency band ratios, including theta/beta, beta/(alpha + beta), theta/(alpha + beta), (theta + alpha)/beta, (theta + alpha)/(alpha + beta), and (gamma + beta)/(sigma + alpha) [14,21–23]. To enhance classification accuracy, some systems integrate additional physiological signals such as electrocardiogram (ECG), EOG, electromyogram (EMG), respiratory activity, or behavioral data [14,23,24]. However, the practical application of these systems remains limited. Most have been validated only under laboratory conditions, typically during early afternoon sessions, after sleep deprivation or not, and their deployment in real-world settings is constrained by the large number of sensors required. To ensure operational feasibility, future development of EEG-based drowsiness detection systems should prioritize the use of a minimal number of sensors - particularly EEG derivations.

To address these limitations, the French company Physip, in collaboration with A Muzet, developed MEEGAWAKE®, an algorithm for detecting and assessing drowsiness, based on the Objective Sleepiness Scale (OSS) framework. The OSS, developed by Muzet et al. [25] was designed to classify sleepiness into five levels based on the duration of alpha and/or theta activity and the presence of slow eye movements, assessed in 20-second epochs. Despite its methodological clarity, the OSS is not widely adopted in subsequent research. Yet a recent study has reported significant associations between OSS scores and sleepiness-related variables, such as sustained attention and driving performance [26], confirming the potentiality of the OSS to offer a good assessment of sleepiness-related cognitive impairments. Whereas the original implementation of the OSS required six EEG and four EOG derivations, limiting its applicability in operational contexts, MEEGAWAKE® algorithm is capable of classifying the five stages of sleepiness identified by the OSS relying exclusively on EEG signals recorded from only two derivations. The performance of this algorithm was validated on 15 subjects in a driving simulator by comparison with visual analysis by a drowsiness reference expert [26]. This streamlined approach facilitates easier integration into workplace environments.

Nevertheless, current real-time spontaneous drowsiness detection systems continue to face several challenges. First, they typically require continuous wear by the operator, which complicates their implementation in real operational settings. Second, the detection of drowsiness often occurs too late to prevent its functional consequences. Therefore, the development of EEG-based tools allowing the prediction of drowsiness and of its behavioral consequences - rather than detecting its presence - would mark a significant advancement in ensuring operator safety. In experimental protocols, routine EEG bio- calibrations, performed with eyes open and eyes closed, are critical for assessing signal quality and characterizing an individual’s resting-state brain activity. Resting-state EEG recordings, under both eyes opened and eyes closed conditions have been used to evaluate task performance and drowsiness levels [27–30]. An algorithm developed by the University of Leipzig (VIGALL) can identify 7 states of somnolence during a 5 minutes resting-state EEG, but requires the recording of at least 19 EEG leads (25 recommended), making it unusable in real operating conditions [31]. These short measures yet offer a promising avenue for prediction: a brief, iterative, and easy to administer resting-state assessment could enable early identification of drowsiness and of its potential cognitive and behavioral impacts, thereby facilitating timely and effective preventive interventions.

The objective of this study is to validate - under sleep deprivation conditions reflecting real-life occupational scenarios (such as delayed, fragmented, or disturbed sleep, typically experienced during night work, shift work, or on-call duty) - the ability of the Objective Sleepiness Scale (OSS) and its automatic analysis by MEEGAWAKE algorithm to accurately assess drowsiness and its associated cognitive and behavioral consequences. In addition, the study aims to evaluate whether automatic analysis of resting-state EEG can reliably predict drowsiness and its functional outcomes, independently of prior wakefulness duration. Such predictive capability could enhance risk assessment and contribute to the prevention of sleepiness-related accidents in operational settings.

We hypothesize that both the OSS and resting-state EEG, potentially adjusted for inter-individual variability in sleepiness susceptibility, can effectively measure and/or predict drowsiness and its behavioral consequences, including the inability to maintain wakefulness and the impairments in sustained attention and vigilance, regardless of the time of day. This approach may provide a robust framework for identifying and mitigating risks associated with drowsiness in safety-critical environments.

## 2. Methods and analysis

### 2.1 Objectives

#### 2.1.1 Main Objective

To determine whether instantaneous drowsiness states, classified either visually using the Objective Sleepiness Scale criteria (OSS) or automatically via the MEEGAWAKE® algorithm, can evaluate the ability to maintain wakefulness, measured by the sleep onset latency during the Maintenance of Wakefulness Test (MWT).

#### 2.1.2 Secondary Objectives

Study’s secondary objectives are:

- to determine whether the instantaneous drowsiness states, classified either visually using the Objective Sleepiness Scale criteria (OSS) or automatically via the MEEGAWAKE® algorithm, can identify the instantaneous functional impairments associated with drowsiness (driving performance, sustained and selective attention, and vigilance).

- to determine whether the instantaneous drowsiness states, classified either visually using the Objective Sleepiness Scale criteria (OSS) or automatically via the MEEGAWAKE® algorithm, can predict the functional impairments associated with drowsiness (wakefulness maintenance, driving performance, sustained and selective attention, and vigilance).

- to validate a robust model, based on a multimodal EEG index derived from resting-state activity recorded during bio-calibration, capable of predicting functional impairments associated with drowsiness, specifically in the domains of wakefulness maintenance, driving performance, sustained and selective attention, and vigilance.

- to identify the determinants of drowsiness, by exploring the effects of individual characteristics (age, sex, chronotype, sleep history, daydreaming tendency) on manifest performance (MWT, cognitive performance) and on introspective sleepiness (assessed by KSS).

### 2.2 Study design

This study is a preliminary in a single-center, non-randomized single-group study performed at the Sleeping department of the SANPSY laboratory, Bordeaux, France.

The recruitment started on March 2023. We expect the recruitment period to last 30 months and to be concluded in September 2025.

### 2.3 Subjects

#### 2.3.1 Sample size calculation

This study is exploratory, since to our knowledge, no other study has used OSS to predict manifest sleepiness. Indeed, no data could allow us to statistically determine the number of subjects required. Consequently, the number of subjects has arbitrarily been set to 40 healthy volunteers sorted in two categories: 28 volunteers (men/women) aged between 20 and 40, and 12 volunteers (men/women) aged between 41 and 60, with a tolerance margin set to 15% for the age category distribution.

#### 2.3.2 Recruitment

Subjects are 40 healthy volunteers recruited from Bordeaux University Hospital’s healthy volunteer database and by advertisement via information-flyers and internet recruitment. Participants will be screened with respect to health status by the medicine doctor, and fill specific questionnaires with professional clinicians. The medicine doctor will provide full information about the study (design, potential risks and constraints, answers to potential questions) before collecting the volunteer’s informed, written, dated and signed consent.

The eligibility criteria include the following inclusion criteria: subjects aged between 20 and 60, with a body-mass index between 18 and 27, having good French skills and ability to understand the study, and being non-professional drivers with valid driver license (obtained at least one year ago). Subjects suffering from severe psychiatric, neurological or medical pathology, or under psychotropic or cardiotropic drug treatments, subjects suffering from chronic insomnia disorder, severe diurnal somnolence or sleeping pathologies that can induce excessive daytime somnolence, subjects declaring substance dependency, alcohol abuse (> 2 glasses/day) and/or excessive consumption of coffee, tea or caffeine-based drinks (such as coke, > 5 cups/day), and subjects that perform night or shift work being on care or on-call duty during the last 72 hours before the experimental sessions, are excluded from the study.

Once included in the study, the participants will answer the following questionnaires, addressing specific items that are exclusion criteria for this study:

- The Patient Health Questionnaire (PHQ-4) is conducted to reveal propensity to anxiety and depression [32]. Subjects are asked to score 4 items reflecting their tendency to feel anxious (2 items) or depressed (2 items) during the previous 2 weeks (on a Likert scale going from 0 : never to 3 : every day). A score above 2 on any item or a total above 4 on anxiety or depression items will lead to an exclusion of the subject.
- The Epworth Sleepiness Scale (ESS) is conducted to reveal daytime sleepiness [33]. The subjects must score their ability to fall asleep in 8 different day-life situations over the last months (on a Likert scale going from 0 : never to 3 : very high probability). Score over 11, reflecting a sever diurnal somnolence, will lead to an exclusion of the subject.
- The STOP-BANG questionnaire scores the Obstructive Sleep Apnea Syndrome’s risk (OSAS) through several items (Snoring, Tiredness, Observed apnea, blood Pressure, BMI, Age, Neck circumference and Gender) [34]. Score over 5, reflecting a high probability to suffer from OSAS, will lead to an exclusion of the subject.
- The Restless Legs Syndrome (RLS) questionnaire scores the risk of suffering from this syndrome through 7 specific questions, to which the subjects must answer (yes/no) [35]. A positive answer to some items, reflecting a Restless Legs Syndrome suspicion will lead to an exclusion of the subject.

After verification of inclusion and exclusion criteria, the experimental sessions will be scheduled. Subjects will undergo 2 experimental sessions, one with a fragmented sleep schedule (3 days) and one with a sleep deprivation schedule (4 days). Each subject’s participation will last 14 to 35 days, according to the timelapse between the 2 experimental sessions, and subjects will receive a financial compensation (900€) for participating to the whole study. Subjects abandoning the study before the end of the protocol will not be considered, and will be replaced, until obtention of 40 healthy volunteers that fulfilled the whole protocol.

### 2.4 Experimental timeline

The experimental schedule is composed of 2 experimental sessions, both designed to increase the sleeping pressure by altering the previous sleep quantity, the first through an imposed fragmented sleep and the second through a full night sleep deprivation. During both sessions, EEG will be recorded (in ambulatory and laboratory conditions) to assess sleep duration and quality, and several tests will be conducted, to assess the effects of sleep alteration on introspective sleepiness (through the Karolinska Sleepiness Scale - KSS), on the capacity to remain awake (through the Maintenance of Wakefulness Test - MWT), on driving performances (driving simulation), on some cognitive functions (sustained attention through the Psychomotor Vigilance Task - PVT, selective attention through the Connor’s Continuous Performance Test II - CPT, and vigilance through the Test of Attentional Performance - TAP) and on the mind-wandering during the tests (through the Conscient Experience Characterization - CEC). EEG will be recorded throughout the tests, to allow drowsiness assessment (through OSS visual and automatic scoring), and resting state EEG will be recorded through regular Bio-Calibrations (BC2 or BC4, respectively lasting 2 or 4 minutes). Details of all the tests and questionnaires are given in the secondary outcome measures section.

#### 2.4.1 Experimental session 1 (ES1) : fragmented sleep

This session lasts 3 consecutive nights and days, during which actimetry and sleep (both ambulatory and in laboratory) are recorded. The 3^rd^ night will be fragmented (with 2 imposed wake-ups, during which attentional tests will be carried out), and during the 3^rd^ day a series of 3 blocks of cognitive and driving tests will be conducted (Fig 2).

**Fig 1.**
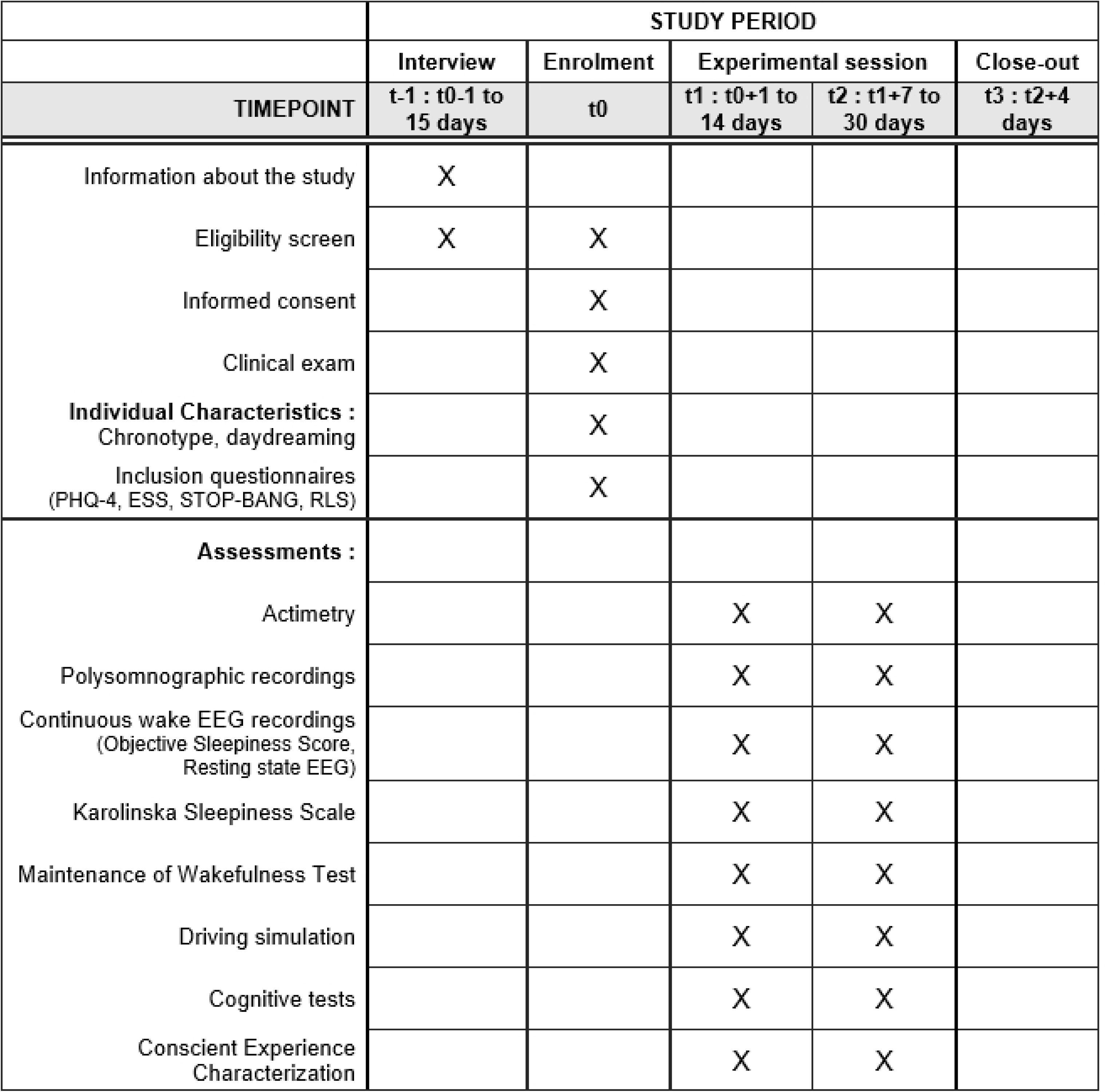
Schedule of enrolment and assessments.

**Fig 2.**
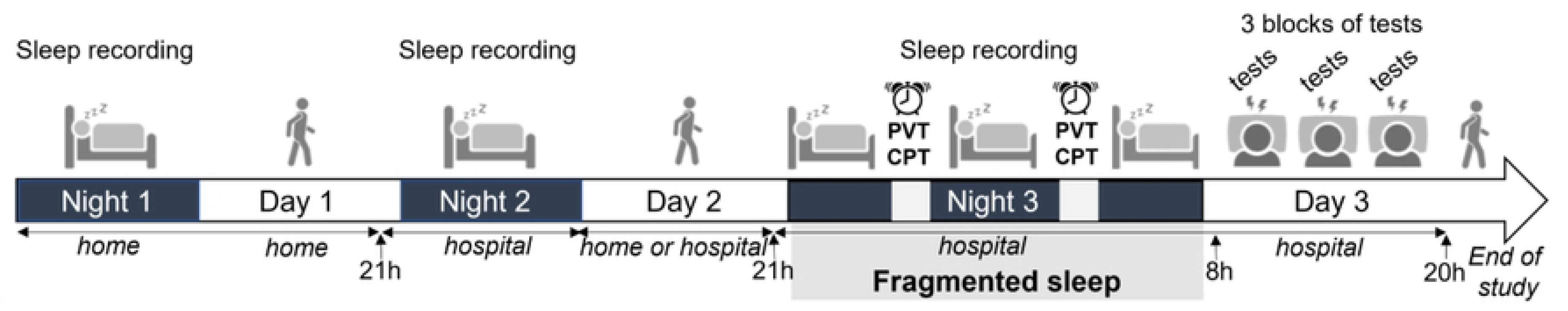
Timeline of ES1: sleep deprivation conditions simulating on-call duty. The dark grey rectangles indicate periods of recorded nocturnal sleep, first at home (Night 1), then in the laboratory for the next two nights (Night 2 and 3). Sleep on Night 3 is interrupted by two half-hour awakenings (light grey rectangles), designed to simulate nocturnal on-call duty, during which the subject performs 2 cognitive tests (CPT and PVT). The following day, the subject undergoes three 4-hour test blocks of cognitive and sleep pressure assessments (grey visual, labeled “tests”, from 8 AM to 8 PM), along with continuous waking EEG recording.

<u>Night 1:</u> In the evening before the 1^st^ night, subjects will come to the laboratory to be equipped with the EMBLA Titanium (recording frequency 256hz, 16-bits resolution) compact ambulatory sleep recorder (single EEG derivation Cz and Pz), to measure their sleep quantity and quality. They will return home for the 1^st^ night, and go to sleep and wake up according to their usual schedule.

Day 1: After waking up and removing the sleep recorder, subjects will carry out their usual activities throughout the day before coming back to the laboratory in the evening, where a training to the cognitive tests and driving simulation will be achieved.

<u>Night 2:</u> Subjects will be equipped with polysomnographic captors (6 derivation EEG : F4, C4, O2, Cz, Pz, A1, 2 EMG, 2 EOG and 2 ECG) connected to Embla NDx amplifier and stored by Natus Sleepworks software (256hz recording frequency, 16-bit resolution) to measure their sleep quantity and quality. They will spend the night in the laboratory, and go to sleep and wake up according to their usual time schedule.

Day 2: After waking up, subjects will either stay in the laboratory or go back to their regular activities before coming back in the evening.

<u>Night 3:</u> Subjects will be re-equipped with polysomnographic captors (as during night 2) and will go to bed no later than midnight. They will be awakened 2 times in the night (2 hours and 4 hours after bedtime) for 30 minutes, during which they will undergo 2 tests to measure their ability to maintain a sustained attention (the PVT and the CPT, detailed in the secondary outcome measures).

<u>Day 3:</u> After waking up at 7am, equipment will be removed and subjects will be equipped with gel-based active electrode system for EEG (Acticap Slim, Brain Products, 8 EEG derivation: Fz, Cz, Pz, C4, O2, C3, O1, P3; 4 EOG vertical and horizontal) connected to a compact wireless EEG amplifiers LiveAmp (Brain Products). The subjects will than undergo three 4h-test blocks, each including a driving simulation, cognitive tests, evaluation of the sleep pressure and of the drowsiness (4-h test block, Fig 3), block 1 will occur from 8-12h, block 2 from 12-16h and block 3 from 16-20h. After the last block, the EEG helmet will be removed and the subject will either stay in the laboratory for a recovery night or get out of the hospital, if accompanied, with recommendation to get home for a recovery night.

**Fig 3.**
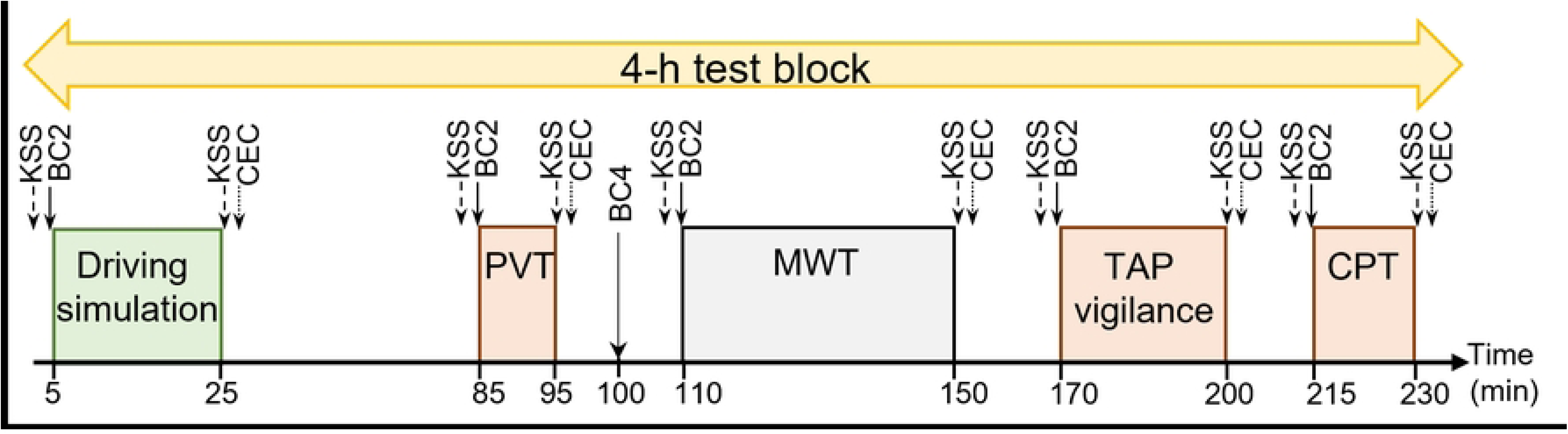
Timeline of the tests and questionnaires over a 4-hour test block. Rectangles indicate the different tests composing the 4-h test block. The first test of the block is a 20min driving session on simulator (driving simulation, minutes 5-25), followed by a 10min reaction time test (PVT, minutes 85-95), a 4min resting state EEG registration (BC4, minutes 100-104), a 40min Maintenance of Wakefulness Test (MWT, minutes 110-150), a 30min vigilance test (TAP vigilance, minutes 170-200) and a 15min sustained attention test (CPT, minutes 215-230). Arrows indicate, before and/or after each test, the registration of the resting state EEG (BC2) and the scoring by the subject of its introspective sleepiness level (KSS) and of its mind wandering during the test (CEC). The pause from minutes 25-85 allows lunch or dinner to be served to the subjects during the relevant blocks (starting at 12 and 8PM).

#### 2.4.2 Experimental Session 2 (ES2): sleep deprivation

This session lasts 4 consecutive nights and 3 days, during which actimetry and sleep (both ambulatory and in laboratory) are recorded. During the 2^nd^ day, subjects will begin a series of six 4-h test blocks, lasting 24h and leading to a full sleep deprivation during the 3^rd^ night. On the 3^rd^ day, subjects will have a morning sleeping recovery time (4h) and begin a new full sleep deprivation with a series of three 4-h test blocks during the 4^th^ night (Fig 4).

**Fig 4.**
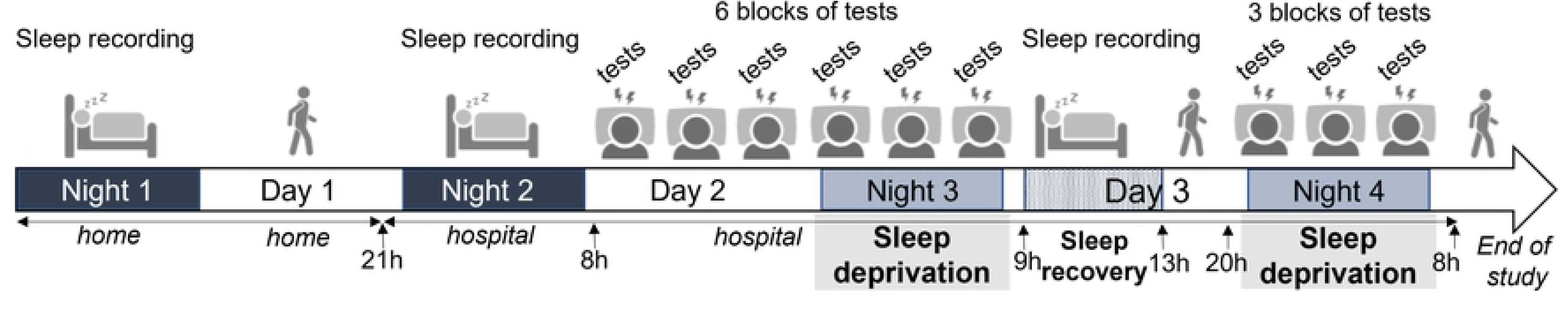
Timeline of ES2: sleep deprivation conditions simulating two successive working nights. The dark grey rectangles represent recorded nocturnal sleep, first at home (Night 1) and then in the laboratory (Night 2). Over the next 24 hours (Day 2 and Night 3, simulating night work), the subject undergoes six 4-hour test blocks of cognitive assessments (grey visual, labeled “tests”, from 8 AM to 8 AM the next day) with continuous waking EEG recording. Light grey rectangles indicate night sleep deprivation (Nights 3 and 4), while the hatched light grey rectangle represents morning recovery sleep. Finally, the subject experiences another night of full sleep deprivation (Night 4), during which they complete three additional 4-hour test blocks of cognitive assessments (from 8 PM to 8 AM) along with continuous waking EEG recording.

<u>Night 1:</u> In the evening before the 1^st^ night, subjects will come to the laboratory to be equipped with the EMBLA Titanium (recording frequency 256hz, 16-bits resolution), compact ambulatory sleep recorder (single EEG derivation Cz and Pz), to measure their sleep quantity and quality. They will return home for the 1^st^ night, and go to sleep and wake up according to their usual schedule.

Day 1: After waking up and removing the sleep recorder, subjects will carry out their usual activities throughout the day before coming back to the laboratory in the evening.

Night 2: Subjects will be equipped with polysomnographic captors (6 derivation EEG : F4, C4, O2, Cz,

Pz, A1, 2 EMG, 2 EOG and 2 ECG) connected to Embla NDx amplifier and stored by Natus Sleepworks software (256hz recording frequency, 16-bit resolution) to measure their sleep quantity and quality. They will spend the night in the laboratory, and will go to bed no later than midnight.

<u>Day 2 and Night 3:</u> After waking up, equipment will be removed and subjects will have breakfast beforebeing equipped with gel-based active electrode system for EEG (Acticap Slim, Brain Products, 8 EEG derivation: Fz, Cz, Pz, C4, O2, C3, O1, P3; 4 EOG vertical and horizontal) connected to a compact wireless EEG amplifiers LiveAmp (Brain Products). The subjects will than undergo six 4h-test blocks (including a driving simulation, cognitive tests, evaluation of the sleep pressure and of the drowsiness, Fig 3). Block 1 will occur from 8-12h, block 2 from 12-16h, block 3 from 16-20h, block 4 from 20h-24h, block 5 from 00-4h and block 6 from 4-8h. After the last block, the EEG helmet will be removed.

<u>Day 3:</u> After the night 3 full sleep deprivation, subjects will be equipped with the polysomnographic captors as described for night 2, to measure their sleep quantity and quality, during a morning recovery sleep (from 9h-13h). After waking up, captors will be removed and subjects will stay in the laboratory without sleeping during the afternoon.

<u>Night 4:</u> Subjects will be re-equipped with gel-based active electrode system for EEG and wireless EEG amplifiers as described in day 2 and night 3 and undergo a new series of three 4h-test blocks (Fig 3). Block 1 will occur from 20h-24h, block 2 from 00-4h and block 3 from 4-8h. After the last block, the EEG helmet will be removed.

Day 4: The subject will either stay in the laboratory for a 5 hours recovery sleep or get out of the hospital, if accompanied, with recommendation to plan a sleep recovery night at home.

#### 2.4.3 Protocol Assistant

Protocol Assistant is a software tool developed by PHYSIP to indicate the timeline of tests and questionnaires over the 4-hour block and to ensure the synchronization with the EEG/EOG acquisition performed by a Brain Products amplifier (e.g. LiveAmp). It is designed to provide event markers (start and end of each test) with a time precision of approximately 1/10th of a second, without requiring any programming skills. Additionally, it allows for the collection and recording of responses to self-assessment questionnaires (KSS and CEC).

After a preliminary configuration phase, during which the timeline for tests and questionnaires is set for the 4-hour block, the software will automatically launch for each acquisition. It will open a graphical interface with windows on 2 separate screens (1 for the experimenter and 1 for the participant), enabling the protocol to be run and the test and questionnaire instructions to be presented to the participant, in written and audio form, while managing the acquisition of EEG signals realized by EEG amplifier.

The software will automatically add time markers to the protocol steps, and enables the experimenter to follow the progress of the protocol via a graphical timeline and via the list of steps, to visually monitor the tracings in real time, to insert customizable predefined event markers, and to interrupt/resume the progress of the protocol.

### 2.5 Outcome measures

#### 2.5.1 Primary Outcome Measure

The Objective Sleepiness Scale (OSS) relies on the analysis of electrophysiological variables to assess the instantaneous drowsiness state, corresponding to the participant’s level of sleepiness. The OSS score, ranging from 0 (full alertness) to 4 (very drowsy), is assigned every 20 second, based on specific EEG activity (amount of beta, alpha, and theta waves) observed simultaneously in two regions of the brain during the epoch, and accompanied by distinct eye blinks and movements (normal or slow). An OSS of 0 corresponds to continuous beta activity, with no alpha or theta activity and no slow eye movement. In contrast, an OSS of 4 is characterized by the presence of alpha and/or theta rhythms for more than 10 cumulative seconds, associated with slow eye movements.

The OSS will be scored both manually, by a trained experimenter - based on EEG (Fz, Cz, Pz, C4, O2, C3, O1 and P3) and EOG (vertical and horizontal) recordings - and automatically, by the MEEGAWAKE® algorithm - based on EEG (Cz and Pz) - every 20 seconds throughout all cognitive tests, to reflect the spontaneous drowsiness states.

The MEEGAWAKE® algorithm uses data-driven criteria to cope with inter-individual variability. After an initial artifact rejection step, the analysis step aims to determine recording-specific thresholds based on EEG power ratios in the usual frequency bands. In addition to this frequency analysis, a temporal localization of EEG events is performed to analyze burst activity. Based on this analysis results, the final step is to classify the 20s-epochs into OSS drowsiness states.

#### 2.5.2 Secondary Outcome Measures

The secondary outcome measures are the following:

The **Maintenance of Wakefulness Test** (MWT) is used to assess the ability to stay awake under sleep- promoting conditions [18,36]. The MWT is a 40-minute trial during which the subject is installed in a comfortable position, in bed, in a quiet dim light room. At the beginning of each trial, the subject is asked to stay awake and fight against sleep, to keep his eyes open and to look straight forward, without looking at the light. During the test, visual analysis of the polysomnographic recording allows to check the subject state, and the test is stopped when unequivocal sleep is observed (defined by 3 consecutive epochs of stage N1 sleep or 1 epoch of any other sleep stage, according to the AASM recommendations [37]. Sleep latency is calculated from defined as the time from lights out until the start of the first epoch of any stage of sleep (an epoch of N1, N2, N3, or R). If the patient succeeds in staying awake during all the test, sleeping latency is set to 40 minutes.

The **resting state EEG** are registered through bio-calibrations, a simple procedure where subjects are placed in a quiet room, asked to relax, look to a picture on the wall, keep their eyes open for 1 minute, refraining from blinking as much as possible, and then close their eyes for 1 minute, without moving (Bio-Calibration lasting 2 minutes - BC2). EEG and EOG channels are recorded continuously during bio-calibrations.

In the preprocessing step of the signal analysis, standard signal processing techniques will be employed, including Notch filtering to remove power-line interference and bandpass filtering (using a Welch filter) to isolate specific frequency sub-bands. The EEG data will be segmented into 4-second epochs to minimize the effects of non-stationarity in longer signal segments. Spectral power across different EEG frequency sub-bands and the frequency gravity center will be computed for each channel under both eyes-open and eyes-closed conditions. To mitigate the influence of outliers, the median spectral power across epochs within each session will be used. EEG signal processing will be performed using the Python MNE toolbox.

Following preprocessing, feature vectors representing each session will be constructed, comprising multiple EEG-derived features. These feature vectors will then serve as inputs for a machine learning model to identify the most informative features for distinguishing between subject’s levels of drowsiness, cognitive, and driving performances as quantified by the different tests conducted after the bio-calibrations.

A longer resting state EEG is registered through a longer bio-calibration, respecting the same procedure as described for BC2, but with subjects asked to keep their eyes open for 2 minutes and then close their eyes for 2 minutes (Bio-Calibration lasting 4 minutes – BC4). The artifact-free Fz signal is then analyzed by 2-second epochs, each of them being characterized by features including the signal magnitude and the spectral power (FFT, Hanning window) in the frequency bands δ (0-4 Hz), θ (4-8 Hz), α (8-12 Hz), σ (12-16 Hz) and β (16-50 Hz). The spectral power of the Fz signal will then be computed in the 6-9Hz band [38] for each 2-second artifact-free epochs. The so-obtained distributions of spectral power values will then be characterized by their mean, standard deviation and median, the mean spectral power being the result of BC4.

The **objective sleep quantity and quality**, measured through polysomnography recordings and analyzed by ASEEGA software (Physip), based on automated analysis of a single EEG channel (Fz-Pz) using a combination of multiple signal processing and classification techniques [39–41]. The preprocessing part aims notably at adapting the analysis criteria to cope with EEG interindividual variability. The analysis part uses both frequential and temporal techniques to retrieve numerous 30-second-long and 1-second-long features as well as performing the detection of several sleep microstructural events. The classification part then reduces and summarizes this information by scoring sleep EEG into conventional sleep stages using artificial intelligence techniques such as pattern recognition and fuzzy logic. This method was validated in healthy individuals and demonstrated high agreement with visual scoring realized by an experiment scorer, between 82.9% and 96% according to the number of vigilance stages considered, allowing its use in several research studies. Classical sleep parameters like the total sleeping time (TST – the sum of all sleep stages, expressed in minutes), the sleep latency (SL – the time, in minutes, from lights out to the first recorded epoch of any sleep stage, N1, N2, N3, or REM), the sleep efficiency (SE - calculated as the ratio of TST to total time in bed, multiplied by 100), the wake after sleep onset (WASO - calculated as the total time in bed minus the TST), and time spent in stage N1, N2, N3 and R (expressed in % of TST and in minutes) will be computed.

The **car driving performances** are evaluated using the fixed-base INRETS-MSIS SIM2 driving simulator [42]. The driving simulator is composed of a computer and a video-game steering wheel with no force feedback applied. The participant’s head is placed at 60 cm in front of the screen. The resolution of the visual scene was 1 024 × 768 pixels and the update rate was 60 Hz. The simulator presents a highway driving scenario on a 19-in. screen. The car’s speed was fixed by the experimenter to 130 km/h and participants were instructed to drive in the center of the right lane. The test lasts 20 minutes, during which the subject must maintain the car’s trajectory on a monotonous two-lane highway (reconstruction of the French A62 highway, without any landscapes or cars). The variability of the car’s lateral position (SDLP) and the number of inappropriate line crossings (ILC) are computed. An ILC was counted each time the car crossed one of the lateral highway lane markers. Standard deviation of the lateral position of the car (SDLP in cm), derived of the vehicle lateral position, indicates weaving of the car.

In healthy subjects, this simulator has been shown to effectively assess nocturnal driving impairment in a dose-response design, considering extended wakefulness and driving duration, compared to real driving conditions [43].

The **cognitive performances** are assessed through 3 standardized cognitive tests:

- the **Psychomotor Vigilance Task** (PVT) is a one-choice serial reaction time task designed to measure the ability to sustain attention [18]. The test was conducted with PC-PVT software, whose performance is comparable to the PVT-192, the gold standard for measuring PVT [44]. The PC-PVT software [45] was installed on a desktop computer running under Windows 10 and equipped with a mouse. The test session lasts 10 minutes, where a visual stimulus randomly appears on a screen, with inter-stimulus intervals varying from 2 to 10 seconds. The subject is asked to react as quickly as possible (by a mouse click) to make the stimulus disappear. Collected data are mean reaction time, slowest and fastest 10 % reaction times and the number of omissions (laps reaction time ≥ 500ms).

- the **Connor’s Continuous Performance Test II** (CPT) evaluates the subject’s capacity to maintain a sustained attention and to discriminate relevant stimulus [46]. The CPT paradigm is based on monitoring and responding to regular nontargets (here, letters), while withholding response to a specific infrequent target (letter “X”). CPT software (MSH, Ver. 5.2) was installed on a desktop computer running under Windows 10 and equipped by a keyboard. The test session lasts 15 minutes, and the subject is asked to push the space bar when any letter is displayed on the screen, except for the letter “X” (target stimulus). Collected data are the mean reaction time (measuring the response execution process) and the number and percentage of omissions (failure to respond to target stimuli, i.e., non-Xs) and commissions errors (responses given to nontargets stimuli, i.e., Xs), which assesses response inhibition.

- the **Vigilance Test of Attentional Performance** (TAP Vigilance), animated line test, measures the ability to focus and to maintain mental effort and vigilance on a monotonous task during an extended period (30 minutes) [47]. The test shows a horizontal line going up and down on a screen with an irregular speed, and from time to time, this line oscillates with a greater amplitude. TAP vigilance (Psytest, Germany) software was installed on a desktop computer running under Windows 10 equipped with a specific external response button. The session lasts 30 minutes and the subject is asked to push on the response button when he notices a difference in the line oscillation amplitude. Collected data are the mean reaction time and the omission (percentage number of missed targets).

The **Karolinska Sleepiness Scale** (KSS) [16] is a subjective scale assessing the level of sleepiness during the past 10 minutes. Subjects must score their ability to maintain wakefulness, on a Likert’s scale, 1, being “perfectly awake” and 9, being “severely somnolent, cannot stay awake”. This score is known to correlate well with EEG measurements and with cognitive and behavioral performances linked to the subject’s ability to maintain wakefulness [48].

The **Conscient Experience Characterization** (CEC) is a subjective scale assessing the level of mind wandering [49]. Subjects are asked to rate their focusing level during the past task, between 5 possibilities

: (a) fully focused (attention and thoughts entirely dedicated to the task) ; (b) focus interference linked to the task (attention and thoughts distracted by task characteristics or by their performance) ; (c) extern distraction (attention and thoughts distracted by environmental stimuli with no relation to the task) ; (d) mind wandering (not focused, with thoughts disconnected from the task or the environment) ; (e) empty mind (not focused, without any particular thoughts).

Finally, the following **individual characteristics** will be assessed:

- the **chronotype** will be assessed by the Horne and Östberg morningness/eveningness questionnaire (MEQ), made of 19 questions, investigating life preferences (activity, wake/sleep cycle, meal), somnolence, and tiredness at certain times of the day [50]. Scores vary from 16 to 86, and allow to identify evening persons (score ≤ 42 for aged 20-44 and ≤ 53 for aged 44-60 subjects), morning persons (score

≥ 58 and ≥ 64 respectively) and extreme chronotypes (score ≤ 31 and ≤ 47 respectively being highly evening persons, and score ≥ 69 whatever the age being highly morning persons; score adaptation to the age of the subject [51]. The French [52] version of Munich Chronotype Questionnaire (MCTQ) [53] will also be used. It is composed of 7 questions investigating sleep habits on working days and 7 questions investigating sleep habits on free days The sleep-corrected local time of mid-sleep on work-free days (MSFsc) and on working days (MSWsc) [53] will be identified and the sleep corrected social jetlag (SJLsc) will be calculated (SJLsc = MSFsc – MSWsc) [54].

- the **Daydreaming Frequency Scale** (DDFS) will be used to assess the subjects’ tendency to daydreaming in everyday situations [55]. Prior to conductions of the experimental sessions, subjects will be asked to score their tendency to daydream during 12 common situations, on a 5-point Likert’s scale (from 1 : very rare, to 5 : very frequent).

### 2.8 Data Analysis

#### 2.8.1 Statistical analysis

The analyses will be performed using specific softwares (R 4.4.2 and IBM SPSS Statistics 27 for Windows). Descriptive statistics and boxplots will be used to explore the data, and the normality of sample distribution (addressed using Shapiro-Wilk test), equality of variance (Levene’s test) and sphericity of the data (Mauchly’s test) will be assessed. We will thus use parametric or non-parametric statistics and consider alpha level α = 0.05.

Multiple correlation between OSS level features, sleepiness (objective and subjective) or cognitive performance will be calculated.

Multivariate analyses of variance (t-tests, Wilcoxon, ANOVA, Kruskall-Wallis or Friedman tests according to the data) will be used and multiple comparisons will be corrected (using Bonferroni correction). Inter-individual data (such as age, gender, daydreaming frequency, chronotype, sleep history) will be introduced as covariates in the models.

We will also construct linear mixed models and multiple and logistic regression models, considering different variables as potential predictors, and conduct analyses of sensitivity and specificity. ROC curve, which visualizes the trade-offs between sensitivity and specificity, will be calculated.

#### 2.8.2 Predictive Models

Given the non-stationary nature of EEG data, machine learning methods capable of handling dataset variability were prioritized. Gradient Boosting was chosen due to its effectiveness in managing mid-sized datasets. Due to the complexity of the model, Explainable Artificial Intelligence (XAI) techniques were employed to provide meaningful insights into the contribution and relative importance of different features in predicting performance levels. Ultimately, this analysis aims to develop a predictive model that can assess performance based on a small subset of highly significant EEG-derived features.

### 2.9 Ethical and safety assessment

This study was approved by the French National Ethics Committee (consultative Committee for the Protection of Persons participating in biomedical research, CPP Sud Est V, on the April 14 2022, under the number N° SI RIPH 2G: 22.00521.000045) and was registered in ClinicalTrials.gov (number NCT05453643). The National Agency for the Safety of Medicines and Health Products was notified about this study. All staff members involved in this study ensure that the full research is conducted in accordance with the ethical dispositions and regulation on research involving human subjects (as stipulated in the Good Clinical Practices (BCP) I.C.H, the law n°2022-323 of March 4, 2022 on research involving human subjects, and the Declaration of Helsinki).

All participants will give their written informed consent prior to inclusion in the study. This study complies with the General Data Protection Regulation (GDPR) and the MR-001 CNIL reference methodology, which require data to be de-identified after collection (for privacy protection). Personal information (such as participants’ names, addresses, medical status) will thus be exclusively managed at the examination center, and not provided to third parties. All data collected will be de-identify, participants being assigned a unique study code, and any information pertaining to personal details is kept in locked filing cabinets in the SANPSY laboratory, only made accessible to authorized research staff directly running the study. Relevant data are entered into an electronic database using only the study codes of each participant, and are securely stored on an encrypted and secure server made accessible to research staff directly running the study through password-protected computers.

Risk of adverse events is considered low, and the study procedure can be cancelled at any time. During the study, participants will be followed by psychologically trained staff, which ensures fast communication of complaints and immediate response. Volunteers will receive a financial compensation (900€) for their participation in the study. The final results of this study will be disseminated through peer-reviewed publications and conferences.

## 3. Discussion

The objective of this study is to validate a new easily implementable spontaneous drowsiness measurement, derived of OSS criteria and based on the automatic analysis of a limited number of EEG channels able to reflect physiological drowsiness and to predict its associated cognitive and behavioral consequences, regardless of the time of day. In addition, the study aims to evaluate whether automatic analysis of OSS or iterative resting-state EEG can reliably predict drowsiness and its functional outcomes, independently of the prior duration of wakefulness. These measurements will enable the early assessment of drowsiness, allowing for the prediction of the ability to maintain wakefulness and/or of both simple and/or complex cognitive impairments associated with drowsiness.

### Effortless installation: The key advantage of our drowsiness detection system

Our EEG-based drowsiness detection system is designed for real-world applications by prioritizing a minimal number of EEG channels and utilizing short, iterative measurements. By reducing hardware complexity and optimizing data collection, our approach ensures practicality in operational conditions, making it suitable for monitoring in real-time scenarios. This not only enhances user comfort and system efficiency but also facilitates seamless integration into sophisticated driver and pilot monitoring systems, contributing to improved safety and performance.

### The Value of 24-Hour EEG Monitoring for the Development of Drowsiness Detection Algorithms

As part of our study protocol, we perform continuous EEG-based monitoring of drowsiness during a 24-hour sleep deprivation period and throughout a subsequent night following daytime recovery sleep. It presents a compelling opportunity to deepen our understanding of the temporal dynamics of sleep pressure and vigilance regulation. Such long-term recordings can capture natural fluctuations in alertness due to circadian and homeostatic processes, as well as individual susceptibility to sleep loss. This comprehensive dataset could inform the development of more robust and generalizable algorithms capable of detecting drowsiness across a wide range of real-world scenarios and time frames

Moreover, a 24-hour EEG assessment allows for the identification of individual variability in drowsiness expression and recovery, potentially enabling personalized predictive models. Such models could dynamically adapt to a user’s physiological state, enhancing the precision and timeliness of drowsiness detection. This approach aligns with the growing emphasis on personalized medicine and human-centered design in neurotechnology.

Developing algorithms trained on 24-hour EEG data may also support the integration of drowsiness monitoring into wearable or minimally invasive systems. By learning to identify EEG markers that are consistent across different levels of activity, posture, and time of day, these algorithms could eventually be embedded in real-time applications—providing continuous monitoring and early warnings to mitigate fatigue-related risks.

### The predictive advantage of our drowsiness detection system based on resting state EEG compared to conventional systems

Unlike conventional automatic drowsiness detection systems that identify spontaneous drowsiness or related performance declines in real-time - often too late to prevent impairments or accidents - our approach predicts drowsiness-induced performance deterioration in advance, enabling timely intervention with effective countermeasures to prevent functional decline

Like certain algorithms, we’ll leverage machine learning analyses while integrating inter-individual characteristics such as age, sex, chronotype, and accumulated sleep deprivation. This personalized approach will enhance predictive accuracy.

By leveraging automatic sleep pressure quantification through resting-state EEG data - adjusted for inter- individual characteristics - our study will provide valuable insights into individual’s vulnerability to drowsiness before critical lapses in attention or vigilance occur. This predictive capability enables proactive interventions, such as fatigue management strategies, to prevent performance deterioration and reduce accident risks.

### Implications for the world of work and transport

The ability to forecast drowsiness-related impairments has significant applications in occupational and transport settings [5,56–58]. Workers in industries requiring night shifts - such as healthcare, emergency response, and manufacturing - frequently suffer from sleep deprivation, which can lead to decreased cognitive performance and increased accident risks during work [5,59,60]. By integrating predictive drowsiness assessment tools, employers can enhance safety measures through “fatigue” monitoring and targeted interventions to prevent fatigue-related errors/accidents.

The transport sector, particularly aviation, rail, and road transport, is highly vulnerable to drowsiness- related accidents [61]. Microsleeps and lapses in vigilance can have catastrophic consequences, making early prediction of drowsiness essential for safety. Unlike real time systems that detect drowsiness only at the moment it occurs, our approach will offer a preemptive strategy by identifying individuals at risk of performance decline related to drowsiness in advance. Our EEG-based drowsiness detection systems can facilitate the implementation of drowsiness-monitoring technologies in pilots, train operators, and long-haul drivers, significantly reducing the likelihood of sleep-related incidents. Additionally, our findings could inform regulatory policies mandating proactive drowsiness risk management in safety-sensitive professions.

Continuous real-time drowsiness assessment is essential, especially for validating Driver Drowsiness Warning systems in Advanced Driver-Assistance Systems (ADAS). To detect spontaneous drowsiness during driving or work scenarios, we will develop an automated algorithm that analyzes two EEG channels in alignment with OSS scoring.

### Clinical Health Applications

Identifying or predicting drowsiness during behavioral or cognitive tasks will primarily be used to assess the effects of medication or of other interventions in conditions related to sleepiness or in patients with excessive daytime sleepiness.

Beyond occupational and transport safety, the study’s findings have important implications for clinical health. Sleep disorders such as insomnia, sleep apnea, and narcolepsy compromise an individual’s ability to maintain wakefulness and cognitive function. By leveraging EEG-based biomarkers to predict drowsiness susceptibility, clinicians could improve diagnostic accuracy and develop more personalized treatment plans. In fact, the MWT is the reference test for measuring manifest sleepiness (ability to maintain wakefulness under monotonous conditions) in sleepy patients. In some countries, MWT has a medico-legal value and is used to decide whether an individual with sleep-related driving risk is fit or unfit to drive after the implementation of therapeutic measures. In this case MWT must be repeated annually for heavy vehicle licenses and every three years for light vehicle licenses. MWT is also recommended to determine the efficacity of treatments against sleepiness. MWT is conducted in specialized sleep centers and requires appropriate equipment and trained personnel for accurate execution and interpretation. However, access to these centers may be limited due to a shortage of specialists and restricted availability of necessary equipment. Automatic analysis of resting-state EEG is simpler, more cost-effective, and easier to administer, making it a potentially valuable alternative to the Maintenance of Wakefulness Test (MWT) for the initial assessment of drowsiness in clinical settings. Automatic analysis of resting-state EEG would be a simpler, more cost-effective, and easier to administer assessment, making it a potentially valuable alternative to the Maintenance of Wakefulness Test (MWT) for the initial assessment of drowsiness in clinical settings. In this sense, the freely available VIGALL algorithm has already demonstrated its ability to provide comparable information on wakefulness regulation as the more resource-intensive and time-consuming Multiple Sleep Latency Test (MSLT) [60]. However, the VIGALL requires 25 EEG derivations, which may still limit its practicality in routine clinical use, as compared to the MEEGAWAKE® algorithm.

Additionally, aged subjects, patients recovering from neurological conditions, such as traumatic brain injuries [62] or neurodegenerative diseases [63,64], often experience disrupted sleep-wake cycles and daytime sleepiness. An easy-to-administer and precise assessment of their ability to stay awake and sustain attention could enhance rehabilitation strategies, improving cognitive recovery and overall well-being.

### Strengths and weaknesses

One of the strengths of this study is its rigorous methodology, aimed at validating a drowsiness detection system through a strictly controlled sleep deprivation protocol, simulating realistic conditions of repeated night work or on-call shifts. The assessment of drowsiness throughout the sleep deprivation periods, both during the day and at night, provides a detailed understanding of its fluctuations according to circadian rhythms and to the accumulation of sleep pressure. The analysis of behavioral and cognitive consequences is particularly thorough, combining subjective and objective measures through simulated driving tests, simple and complex cognitive tasks, and a scientifically recognized test for evaluating the ability to stay awake. Additionally, the sample is balanced in terms of gender and covers a wide age range (18–65 years), enhancing the generalizability of the results to a healthy adult population.

However, this study also has certain limitations. Although the laboratory setting minimizes environmental biases and ensures strict standardization of experimental conditions, it does not fully replicate the constraints and dynamics of real-world settings, particularly in professional environments. Furthermore, the relatively small sample size (n=40) limits the statistical power of the conclusions. Another notable limitation is the exclusion of patients suffering from sleep disorders or excessive drowsiness, restricting the applicability of the findings to healthy individuals only. As a result, it is not possible to confirm that the tested system would be equally effective in a clinical population. These limitations should be considered before broader implementation of this technology.

## 4. Conclusion

By demonstrating that automatic analysis of OSS and resting-state EEG parameters can predict drowsiness and its mid-term behavioral consequences, this study will establish a foundation for proactive drowsiness management strategies in occupational, transport, and clinical settings. Unlike real-time drowsiness detection systems, which may be too late to prevent accidents, our approach allows for timely interventions that mitigate risks before performance declines become hazardous. The ability to forecast drowsiness-related impairments has far-reaching benefits, from reducing workplace and transport accidents to improving clinical care and patient outcomes. Future research should aim to validate these findings across diverse populations and operational conditions, ensuring the widespread implementation of this drowsiness detections in the management of accidental sleep-related risks.

## Data Availability

No datasets were generated or analysed during the current study. All relevant deidentified data will be made available upon study completion and results publication.

## Acknowledgments

The Bordeaux University, the French National Centre for Scientific Research (CNRS) and the staff of PRNPP facility and his leader Pierre Philip for their support in the implementation of this study.

